# Network analysis shows decreased ipsilesional structural connectivity in glioma patients

**DOI:** 10.1101/2021.06.22.21259319

**Authors:** Lucius S. Fekonja, Ziqian Wang, Alberto Cacciola, Timo Roine, D. Baran Aydogan, Darius Mewes, Sebastian Vellmer, Peter Vajkoczy, Thomas Picht

## Abstract

Gliomas that infiltrate networks and systems, such as the motor system, often lead to substantial functional impairment in multiple systems. Network-based statistics (NBS) allow to assess local network differences (1) and graph theoretical analyses (2) enable investigation of global and local network properties. Here, we used network measures to characterize glioma-related decreases in structural connectivity by comparing the ipsi- with the contralesional hemispheres of patients and correlated findings with neurological assessment.

We found that lesion location resulted in differential impairment of both short and long connectivity patterns. Network analysis showed reduced global and local efficiency in the ipsilesional hemisphere compared to the contralesional hemispheric networks. In network science, reduced global and local efficiency reflect the impairment of information transfer across different regions of a network.

**One-Sentence Summary:** Network analyses show glioma-related decreased ipsilesional structural connectivity and correlates with neurological status.

## Introduction

Brain tumors often lead to significant functional impairments that are not always linked to the specific localization of the tumor. While classical theory assumes that local lesions have an exclusively local impact, there is increasing evidence that brain tumors affect the entire anatomical network of the brain (*3*). Therefore, mapping the effects of brain tumors on spatially distributed neuronal networks may be essential for a better understanding of anatomical-functional relationships (*4*). In recent years, network neuroscience has contributed significantly to mapping brain function to structure and has advanced precision medicine by identifying quantitative biomarkers for assessing brain disease severity (*5*). Toolboxes for network-based analyses, such as Network-Based Statistics (NBS), allow connectome-wide non-parametric analyses to identify groups of connections showing a significant effect while controlling the family-wise errors (*1*). Although NBS does not provide information on network topology, complex-network measures of global- and local-scale network organization (*2, 6*) have emerged as valuable and reproducible tools for exploiting the topological network architecture of the brain in healthy and diseased subjects (*7, 8*). Graph theoretical network analysis, applied to structural and functional connectome data in glioma patients, has already shown that network efficiency correlated with cognitive performance in IDH1 wildtype astrocytoma (*9*) and that alterations in distinct connectome profiles are related to clinical phenotype in newly diagnosed glioma patients (*10*).

Here, we combine tractography with graph theoretical analysis and NBS to assess tumor-related structural connectome alterations within the ipsilesional hemisphere of glioma patients. In addition, to address the variability in results due to the choice of tractography algorithm, we employed two different algorithms: (i) deterministic and (ii) probabilistic. Moreover, in contrast to many other network analysis studies (*11*), we used a state-of-the-art diffusion MRI processing pipeline that involves constrained spherical deconvolution (CSD) for the estimation of fiber orientation distributions (FODs) (*12*), anatomically constrained tractography (*13*) and spherical-deconvolution informed filtering of tractograms (*14*) within the MRtrix3 open software work frame (*15*). The use of different tractography algorithms allowed us to better compare and interpret the results. With this study, we aim to gain detailed insights into how World Health Organization (WHO) grade II-IV gliomas affect cerebral networks and lead to altered connectivity patterns that affect motor and non-motor functions. We hypothesize that asymmetries between ipsi- and contralesional connectivity profiles are related to specific tumor locations that correlate with functional impairment or neurological patient status.

## Results

The assessment of MRC, NIHSS and tumor volume and TMS mapping related RMT determination was feasible in all patients. NIHSS was discarded in three patients due to incomplete data. Enantiomorphic lesion filling, FastSurfer segmentation, iFOD2-based and SD_STREAM-based connectome construction was possible in each subject.

### Network based statistics

We obtained 30 iFOD2-related and 19 SD_STREAM-related significant edges by TFNBS on the entire cohort (Fig. 2). Furthermore, we obtained 15 iFOD2-related and 14 SD_STREAM-related significant edges in the precentral subgroup and 15 iFOD2-related and 6 SD_STREAM-related significant edges in the insular subgroup (Fig. 2). Frontal and postcentral groups revealed no significant differences by TFNBS.

**Figure 1:**
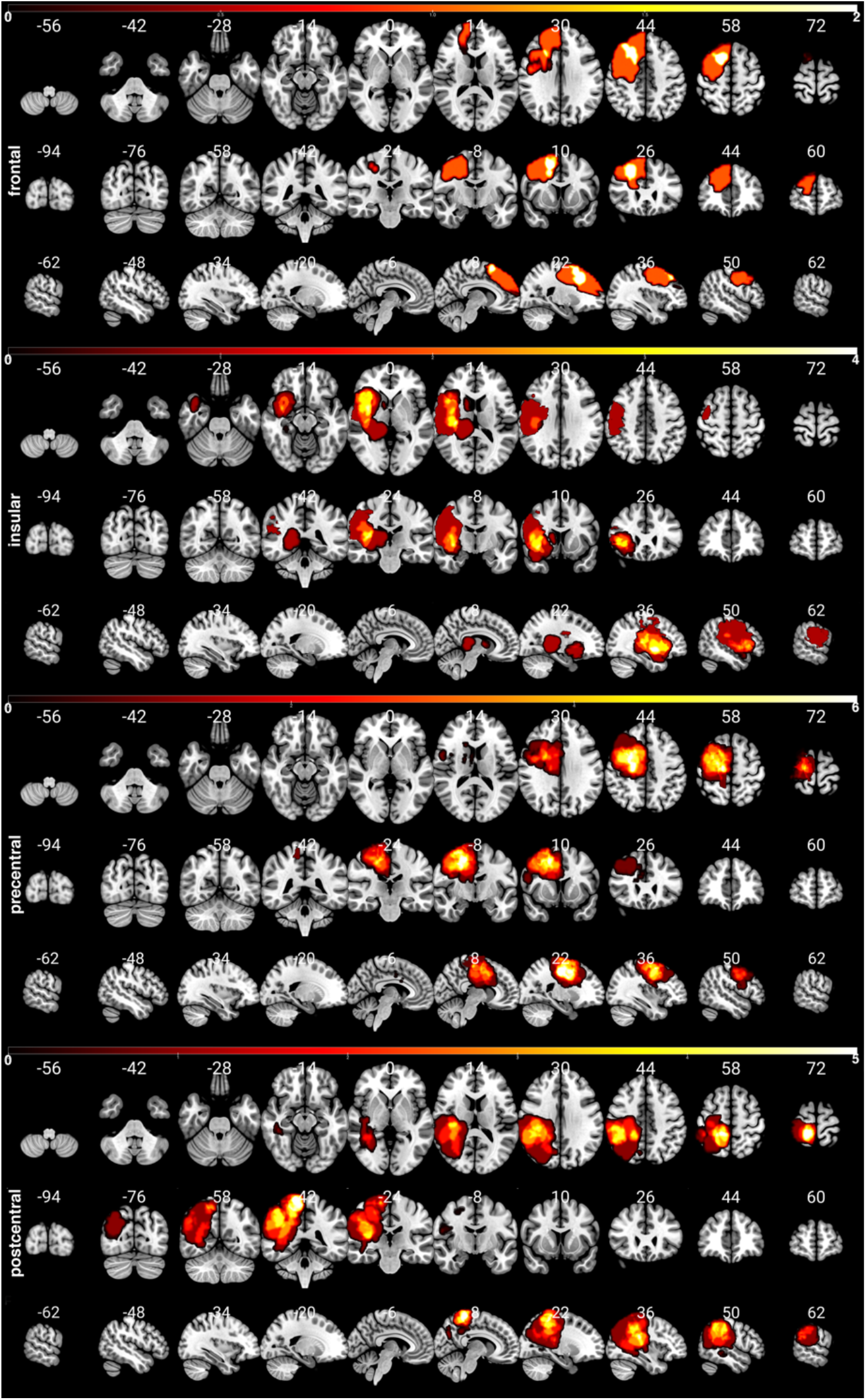
Distribution of the subgroups’ patients’ lesions (frontal, insular, precentral and postcentral). The color bar indicates the occurrence of lesions per voxel. To enable a clear comparison of lesion location, lesions of the left hemisphere were mapped to the right hemisphere.

**Figure 2:**
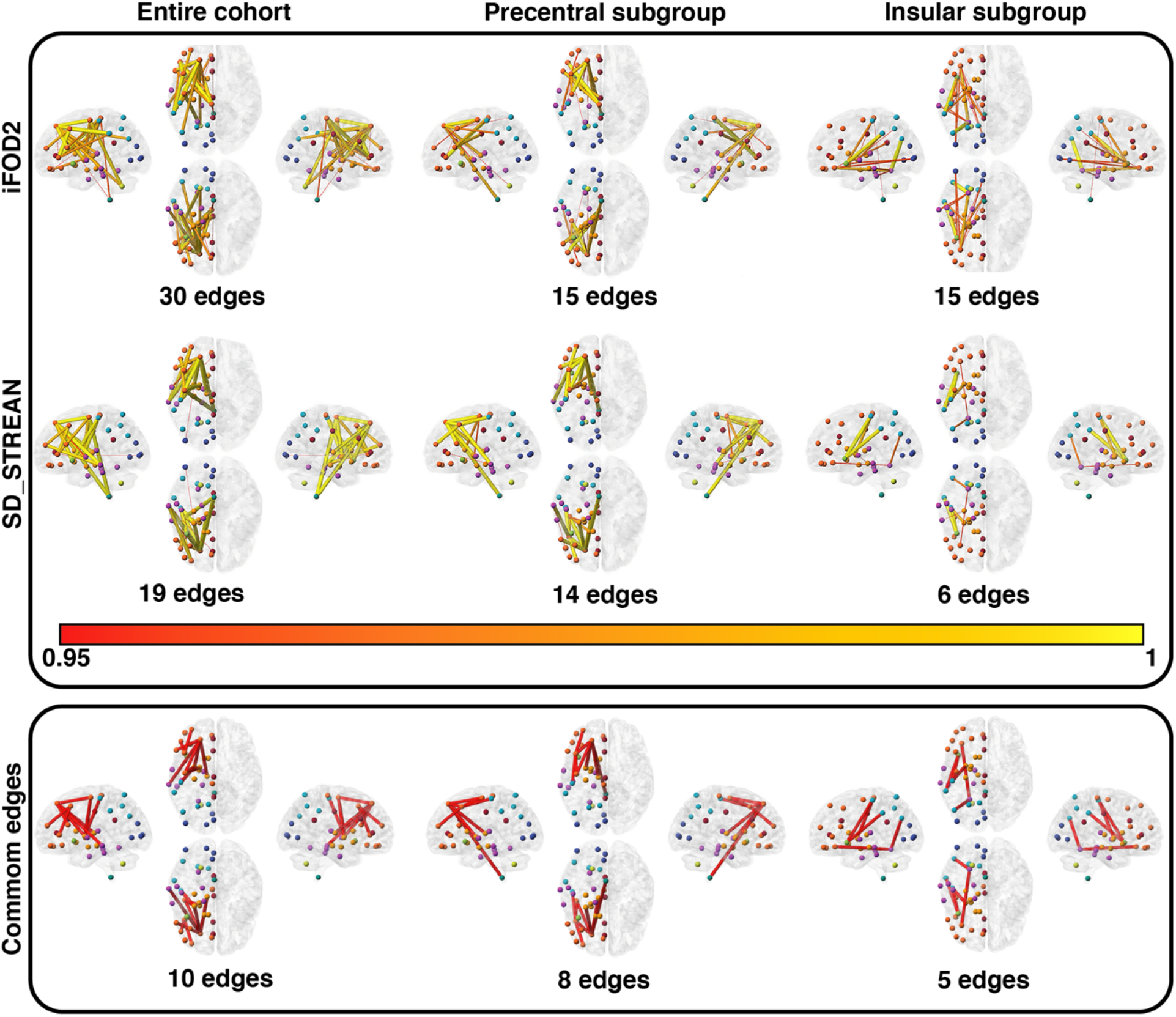
TFNBS results. The heatmap indicates the FWE-corrected significance (1-p, only significant edges are displayed with p-values < 0.05). Upper box, top row: iFOD2-based analyses (left=entire cohort, middle=precentral group, right=insular group). Middle row: SD_STREAM-based analysis (left=entire cohort, middle= precentral group, right= insular group). Lower box: Significant edges that were significant in both tractography algorithms (p-values < 0.05), cf. supplementary table 3.

We performed Spearman correlation analyses between the strength of the edges revealed as significant by TFNBS and MRC, NIHSS, RMT ratio, tumor volume and WHO degree variables for ipsilesional, contralesional and differences between ipsi- and contralesional matrices. The Spearman correlation analyses resulted in 6 (3 iFOD2-based, 3 SD_STREAM-based) FDR-corrected significant relationships (Fig. 3).

**Figure 3:**
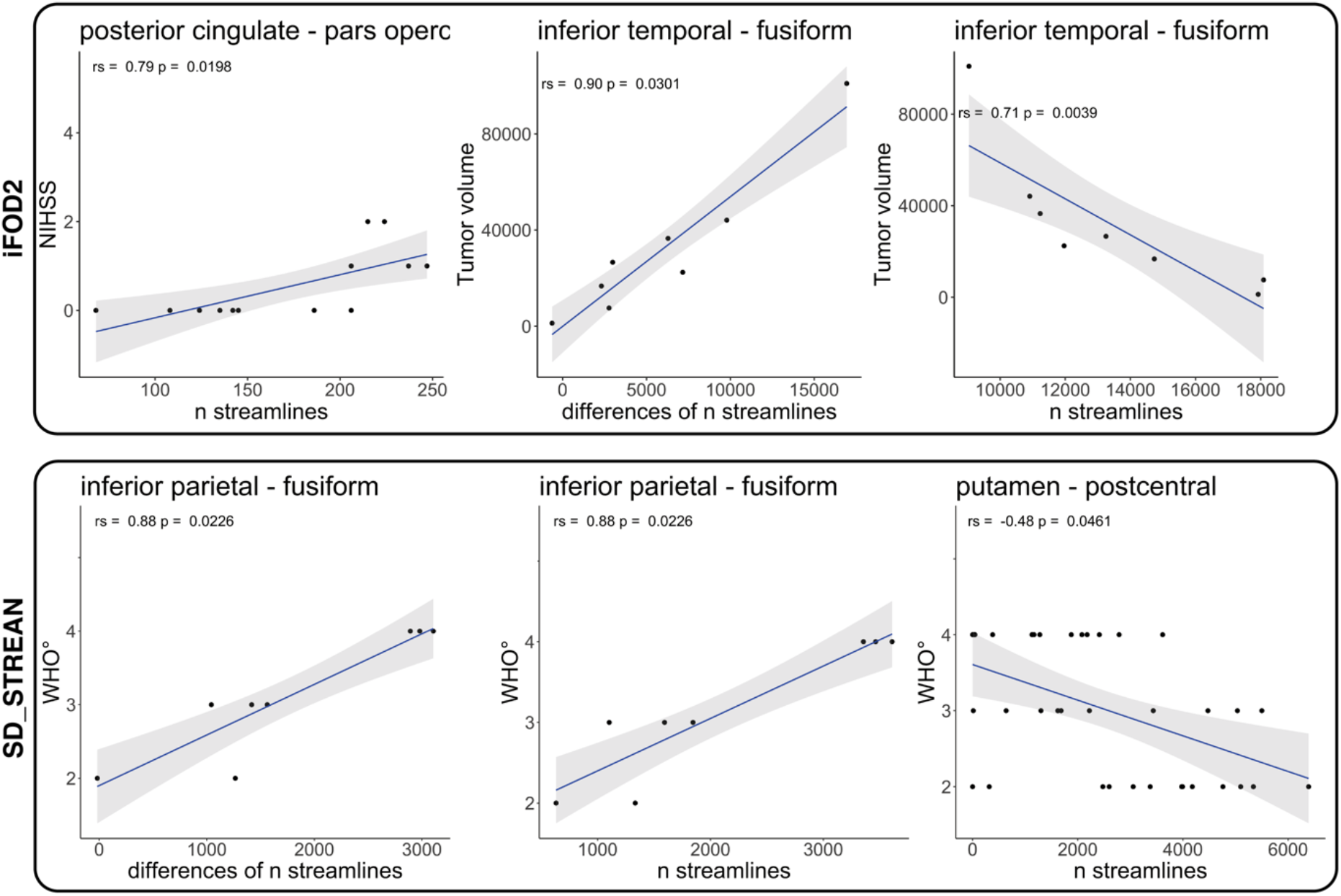
Line plots of FDR-corrected Spearman significant correlations between TFNBS-selected significant edges and NIHSS, WHO degree or tumor volume.

### Probabilistic tractography

Based on iFOD2 connectome matrices, there was a significant positive correlation between the streamlines’ strength of posterior cingulate - pars opercularis, and NIHSS in the contralesional hemispheres and precentral group, rs(13) = .79, p = .0238, a positive correlation of tumor volume and inferior temporal gyrus - fusiform gyrus, rs(8) = .90, p = .0301 regarding the differences of ipsi- and contralesional matrices in the insular group and a negative correlation of tumor volume and inferior temporal gyrus – fusiform gyrus, rs(8) = -.71, p = .0039 in the ipsilesional insular group.

### Deterministic tractography

Based on SD_STREAM connectome matrices, there was a significant positive correlation between the weight of inferior parietal gyrus - fusiform gyrus and WHO degree in the difference between contra- and ipsilesional hemispheres and insular group, rs(8) = .88, p = .0226 as well as a significant positive correlation between the streamlines strength of inferior parietal gyrus and fusiform gyrus and WHO degree in the contralesional hemispheres and insular group, rs(8) = .88, p= .0226. Furthermore, we observed a significant negative correlation between the streamlines strength of putamen-postcentral gyrus and WHO degree in the ipsilesional hemispheres and the entire cohort, rs(37) = -.48, p = .0461.

### Complex network analysis

Graph theoretical analysis showed significant correlations with RMT ratio, small worldness and local efficiency in contralesional hemispheres (rs(37) = -.373, p = .0228), but not with any clinical measure (i.e., MRC, NIHSS, WHO degree, tumor volume) for deterministic and probabilistic tractography results. There were no significant differences for assortativity, nodal degree, hierarchy, nodal efficiency, rich club and small worldness between contra- and ipsilesional hemispheres. However, for some global network metrics, there were significant differences between contra- and ipsilesional hemispheres (see below). iFOD2-based differences resulted in a correlation of MRC with hierarchy (rs(37) = 0.372, p = 0.02339) and local efficiency (rs(37) = 0.349, p = 0.03432). SD_STREAM-based differences resulted in a correlation of MRC and local efficiency (rs(37) = 0.326, p = 0.0489) and NIHSS and local efficiency (rs(37) = -0.361, p = 0.03606), cf. Fig. 4.

**Figure 4:**
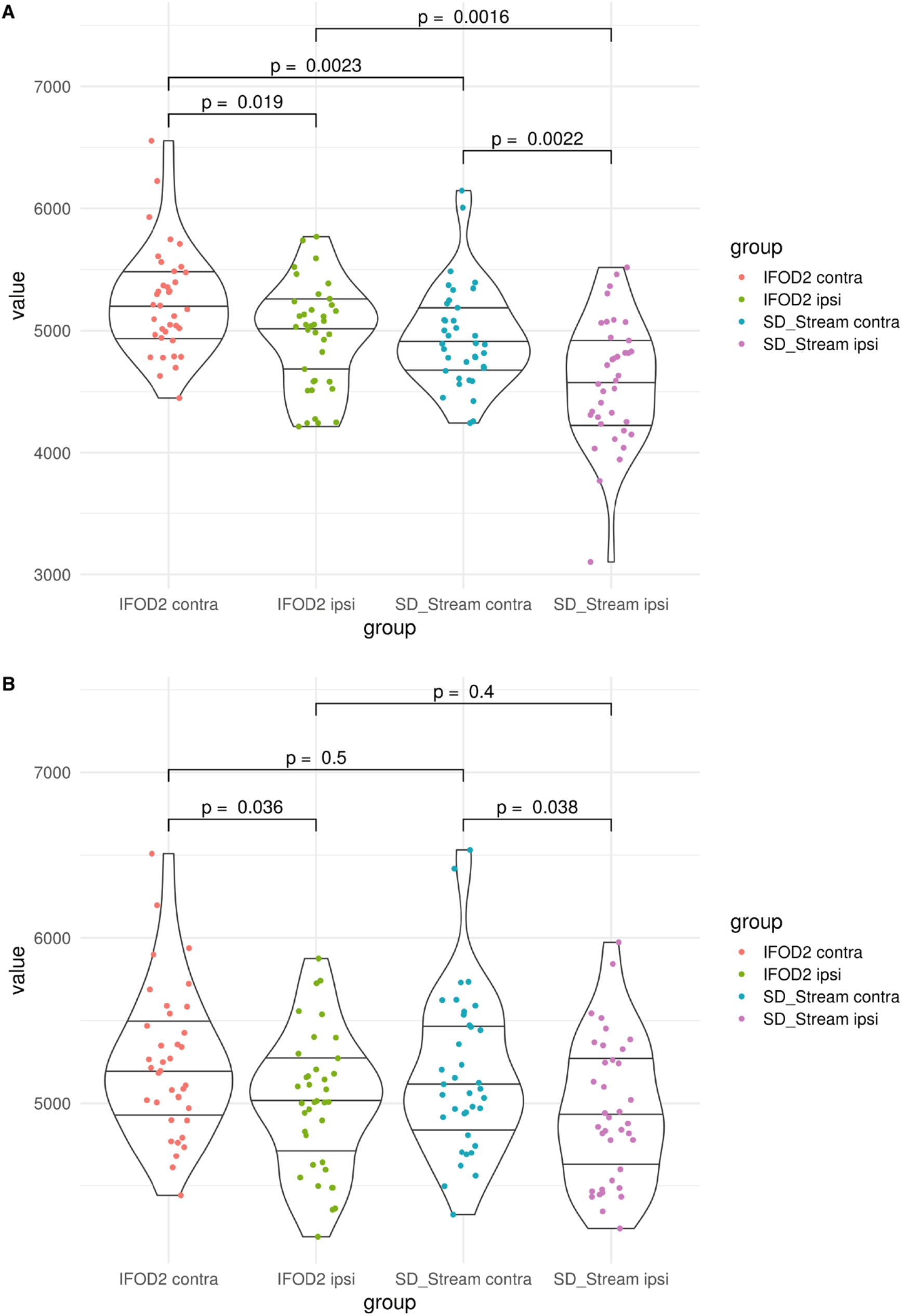
Violin plots of Wilcoxon tests for global (A) and local efficiency (B) by testing the differences of ipsilesional (*_ipsi) and contralesional (*_contra) hemispheres in relation to the tractography algorithms.

### Probabilistic tractography

Based on iFOD2 connectome matrices, there was a significant decrease in the global efficiency of ipsilesional hemispheres (M = 4955, SD = 429) compared to the contralesional hemispheres (M = 5239, SD = 436), t(36) = 3.3, p = .0024 as well as a significant decrease in the local efficiency of pathological hemispheres (M = 5006, SD = 411) compared to the healthy hemispheres (M = 5241, SD = 446), t(36) = 3.05, p = .0043, cf. Fig. 4.

### Deterministic tractography

Regarding SD_STREAM-based connectome matrices, there was a significant decrease in the global efficiency of ipsilesional hemispheres (M = 4584, SD = 507) compared to the contralesional hemispheres (M = 4950, SD = 415), t(36) = 3.7, p = .00078 and a significant decrease in the local efficiency of ipsilesional hemispheres (M = 4948, SD = 434) compared to the healthy hemispheres (M = 5176, SD = 482), t(36) = 3.34, p = .0019, cf. Fig .4. Furthermore, SD_STREAM and iFOD2 matrices were found to be strongly positively correlated in local and global efficiency in ipsi- and contralesional groups. Global efficiency contralesional: *rs*(72) = *0*.*92, p* < .000, *Global ipsilesional: rs(72) = (0*.*86, p <* .*000, local efficiency contralesional: rs(72) = 0*.*93, p <* .*000 and local efficiency ipsilesional*: rs(72) = *0*.*88, p =* .*000*. In addition to the correlations, local efficiency showed no differences between the two tractography algorithms with respect to the same hemisphere, but significant differences between ipsi- and contralesional hemispheres appeared, whereas global efficiency showed differences between the two hemispheres as well as between the two tractography algorithms with respect to the same hemisphere (cf. Fig. 4).

## Discussion

The last decades have been marked by significant advances in the characterization of brain morphology, function and brain disorders using connectomics (*56*). The localizationist theory has been assimilated into associationist models that see the brain organized in parallel distributed networks (*3*). Primary sensory and motor functions are considered more focally localized (*57, 58*), while higher cognitive functions are discussed as organized in large-scale (*59*) distributed networks. Brain functions and complex behavior are thought to arise from parallel processing and integration performed by large-scale distributed networks rather than single epicenters (*60-62*). In this scenario, connectomics and graph theory could prove as powerful tools to map and relate the architecture of brain networks from structure to function and to identify specific neural substrates associated with dysfunction (*6, 63*).

We constructed structural connectomes from glioma patients to investigate the effects of tumors on ipsi- and contralesional networks. To provide a comprehensive overview and better characterize our results, we used two different tractography algorithms, namely the probabilistic CSD-based iFOD2 and the deterministic CSD-based SD_STREAM, which resulted in two distinct types of structural connectomes.

Although several studies have shown that tumor-induced changes in the WM could be analyzed using dMRI-based methods (*11, 64*), to our knowledge, this is the first study characterizing quantitatively and qualitatively structural connectivity changes in tumor patients at the hemispheric network level.

### Glioma-related hemispheric differences

An interesting finding of this study is the impairment of the ipsilesional structural connectivity compared to the contralesional hemisphere. Earlier studies have shown an increase in ipsilesional raw fiber count (*65*), as well as an increased functional connectivity of the hippocampus and antero-medial portion of the posterior cingulate in glioma patients compared to healthy controls (*66*). However, from a structural connectivity perspective, these findings are likely to be more widely present than the morphological effects of glioma, including tract displacement, edema, blood-brain-barrier disruption, necrosis, or degradation of the surrounding cerebral tissue (*67, 68*). In line with this hypothesis, TFNBS analysis identified significant disconnected subnetworks involving fronto-frontal, fronto-parietal, and fronto-insular edges in the ipsilesional hemispheres compared to the contralateral ones. Furthermore, we observed a significant negative correlation between streamline strength of the putamen-postcentral gyrus connectivity in the ipsilesional hemisphere and WHO degree in the entire cohort. This result suggests that higher WHO grade gliomas are more likely to impair the cortico-subcortical connections regardless of their location, thus further reinforcing the idea that patterns of altered structural connectivity depend on the WHO grade (*10, 69*).

As earlier studies have demonstrated local effects of gliomas on their surrounding microstructural integrity of the WM (*70, 71*), we aimed at investigating whether glioma locations may affect unique subsets of nodes and edges in the ipsilesional network. While gliomas affect brain regions and connections near the lesion, brain connectivity changes may also occur distally from the lesion, either because of the resulting local dysconnectivity or as a neuroplasticity mechanism taking place during tumor growth. In line with this hypothesis, our subgroup analysis showed that lesion location (for instance, pre-central and insular) affected connectivity patterns that belong to distinct subnetworks. We obtained several iFOD2- and SD_STREAM-related significantly affected edges in the precentral and in the insular subgroup. By contrast, no significant TFNBS differences emerged for the frontal and postcentral groups, neither using iFOD2-nor SD_STREAM-based connectome matrices. Additionally, we found a significant positive correlation between streamline strength of the posterior cingulate - pars opercularis edge of the contralesional hemisphere and NIHSS in the precentral subgroup, as well as a significant positive correlation between streamline strength of inferior parietal gyrus - fusiform gyrus and WHO degree in the contralesional hemisphere in the insular subgroup. These findings may suggest that the presence of increased structural connectivity in the contralesional hemisphere is correlated both with worse clinical conditions and higher-grade gliomas, suggesting possible compensatory adaptive changes of connectomic profiles in contralesional networks in patients with unilateral gliomas (*10, 72*). In addition, we found that tumor volume was correlated with the inferior temporal gyrus-fusiform gyrus edge. A larger tumor volume in the insular region likely impairs the connection strength in the ipsilesional hemisphere in this patient subgroup.

Furthermore, the fusiform gyrus, particularly its anterior portion, has been shown to be strongly connected to the inferior temporal gyrus, which, as part of the human ventral visual cortex, plays a role in higher-order visual processing such as face perception and object recognition (*73, 74*). Overall, our results not only demonstrate impaired ipsilesional structural connectivity in glioma patients, but also suggest that a possible neural basis of higher-order symptoms may occur due to the distributed organization of function.

### Network-based analysis

TFNBS offers high sensitivity in detecting altered patterns of connectivity in a network. However, TFNBS data are not specific to any network topology measure, i.e., they cannot offer information related with a particular property of the topology that differs between the hemispheres, despite the identified sub-networks displaying significant between-hemispheres differences. Therefore, the deeper understanding of brain topology and the extent to which a network holds certain topological characteristics, e.g., integration and differentiation, are important to find key elements supporting the structural connectivity involvement in glioma patients. About that, complex network topology analysis of the structural connectomes allows for investigating both the global and local topological organizations, as well as specific connections between the regions. Hence, to provide a comprehensive characterization of the hemispheric structural network changes, we computed especially two common network measures, namely global and local efficiency. Both iFOD2-based and SD_STREAM-based structural connectomes showed reduced global and local efficiency in the ipsilesional hemisphere compared to the contralesional hemispheric networks. Global efficiency is a measure that is inversely related to the topological distance between nodes and quantifies how efficiently the information is exchanged within the network. The reduced values of global efficiency suggest less efficient pathways from one brain area to another and, consequently, lower levels of integration in the ipsilesional hemispheric network.

The average local efficiency quantifies the ability of fault tolerance of the network measuring the information exchange of the subnetwork consisting of itself and its direct neighbors. Therefore, the lower values of local efficiency found in the ipsilesional hemispheric network suggest that the structural brain network of glioma patients is topologically organized to minimize segregation of neural processing.

Reduced global and local efficiency reflect the impairment of information transfer across different regions of derived networks, which is likely linked to the pathological involvement of both long- and short-range connections. In contrast with these findings, a recent publication showed increased global and local efficiency in the ipsilesional hemispheric network, identifying unique sets of nodes with changes in network efficiency depending on lesion location (*71*).

### Tractography-algorithm relationships

As tractography has been discussed to provide high rates of false-positive (Thomas et al. PNAS 2014, Schilling et al. NeuroImage 2019) and false negative (Aydogan et al. Brain Structure and Function 2018) streamlines and no gold-standard approaches have been identified in general, we used two different tractography algorithms to confirm our findings – probabilistic CSD-based iFOD2 and deterministic CSD-based SD_STREAM. Both algorithms identified the involvement of similar, ipsilesional decreased structural connectivity patterns, with the only difference in the resulting number of decreased edges, confirming the plausible anatomical reliability of our findings (cf. Figure 2). Here, we found SD_STREAM and iFOD2 to be strongly positively correlated in local and global efficiency both in ipsi- and contralesional hemispheres, however, the absolute values of these measures were found to be significantly different between the two approaches. Indeed, despite probabilistic tractography approaches have been shown to outperform deterministic tracking in reaching the full extent of the bundles (*75*), the latter are still widely used in clinical settings, and moreover they are used based on diffusion tensor imaging (DTI). Deterministic tractography algorithms proceed by stepping along the principal direction of diffusion, and thus they do not address uncertainty in fiber orientation. CSD-based probabilistic algorithms on the other hand, assume a distribution of possible orientations for propagation taking into account uncertainty in streamline segment orientation. While this may result in a larger number of false-positive streamlines (with relatively low streamline density), probabilistic algorithms can identify tract segments that are not reconstructed by the deterministic approach, thus potentially reducing the risk of underestimating the real extent of the tracts, i.e., reducing false-negatives (*75*). Finally, with regards to connectomics and graph theory, our findings of a linear positive correlation between network measures derived by deterministic and probabilistic CSD-based tractography are in line with previous studies showing significant correlations between link-wise intraclass correlation coefficients from both methodologies (*76*). Altogether, while our deterministic and probabilistic tractography based TFNBS and network measures findings have noticeable differences, they also suggest a certain level of consistency in anatomical reproducibility. Importantly, we did not find significant differences in local efficiency with respect to the two tractography algorithms (cf. Figure 4), that further strengthens our finding of decreased local efficiency in the ipsilisional hemisphere.

#### Clinical correlations

Available data in the literature left the problem, whether a correlation between clinical scores, histopathology, RMT and network topology measures exists, partially unsolved. Even though global and local efficiency provided a between-hemispheric differentiation, NIHSS, WHO grade, RMT values were poorly correlated. This may be related to the fact that macro- and microscopic brain damage, as in glioma patients, possibly leads to a comparable impairment of network measures in terms of connectivity strength reduction and reorganization phenomena throughout the ipsilesional hemisphere.

### Limitations

FastSurfer is not trained with images generated with enantiomorphic filling and thus might have performed differently to using healthy subject data as input. However, FastSurfer employs reliable landmarks while parcellating brain images and has been shown to outperform FreeSurfer with respect to runtime, test-retest reliability, and sensitivity. Tractography methods suffer from a range of limitations that make its routine use problematic. It is well known that tractograms contain false positive (*77*) and false negative (*78*) streamlines (*79*)In addition, tractography cannot distinguish between afferent and efferent connections, and streamlines may terminate improperly (*80*). Furthermore, the use of SIFT in pathological connectomes is currently being discussed (*46, 81-83*), however, we did not observe any disadvantages. The dMRI data used for this study consists of a typical clinical single-shell acquisition, and is thus suboptimal due to incomplete attenuation of apparent extra-axonal signal (*84*). Furthermore, our results are atlas dependent and other atlas choices would result in different parcellations and edge assignments to nodes (*2*).

Moreover, structural brain asymmetries appear to be present even in healthy subjects. A study regarding structural network topology showed that the right hemisphere network is less efficient than that of the left hemisphere (*49*). Specifically, the right hemisphere showed higher values of betweenness centrality and small-worldness, indicating a less optimal organization for information processing and a more random configuration compared with left-hemispheric networks. In contrast, a further study found higher global efficiency in the right hemisphere compared with the left hemispheric network (*85*). Finally, the small sample sizes for insular (n=8) and frontal (n=3) lobe subgroups make these data susceptible to both outliers and false negatives.

### Conclusions

In the present study, we showed altered ipsilesional connectivity in patients with unilateral gliomas. TFNBS analysis identified significant disconnected subnetworks involving fronto-frontal and fronto-insular connections in the ipsilesional hemisphere compared to the contralateral one. Our subgroup analysis also showed that the lesion location (e.g., pre-central and insular) affected connectivity patterns which belong to distinct and peculiar subnetworks, thus highlighting the pivotal role of lesion location in driving corresponding connectivity changes. Such connectivity changes were accompanied by reduced global and local efficiency of the ipsilesional network, suggesting tumor-related altered information transfer, which is due to the pathological involvement of both long- and short-range connectivity patterns and disturbed network integration. Moreover, we observed a correlation between the difference of the matrices in terms of hierarchy as well as local efficiency and functional impairment scales, such as MRC and NIHSS. Additionally, deterministic and probabilistic connectome matrices correlated strongly positively in local and global efficiency in ipsi- and contralesional groups. The integration of connectomics into clinical applications is of paramount importance and provides a novel perspective in the neurooncological scenario having the potential to revolutionize personalized medicine and therapy. Indeed, studying structural connectivity in brain tumor patients through the lens of network neuroscience may guide and improve tumor resection while preserving important nodes and edges which are located more distant from the lesion but also involved in motor and cognitive functions. Finally, network neuroscience represents an important computational approach to better understand glioma induced structural changes and contributes to our understanding on the relationship of network topology to motor function.

## Data Availability

The datasets and scripts used in this manuscript are
available at: https://github.com/CUB-IGL/Network-analyses-reveal-global-and-local-gliomarelated-
decreases-in-ipsilesional-structural-connect

https://github.com/CUB-IGL/Network-analyses-reveal-global-and-local-glioma-related-decreases-in-ipsilesional-structural-connect

## Acknowledgments

LF, DM and TP acknowledge the support of the Cluster of Excellence Matters of Activity. Image Space Material funded by the Deutsche Forschungsgemeinschaft (DFG, German Research Foundation) under Germany’s Excellence Strategy – EXC 2025 – 390648296.

TR received support from the Finnish Cultural Foundation. Figure 2 was visualized with the BrainNet Viewer toolbox (86).

## Author contributions

Conceptualization: L.S.F., A.C., T.R., B.A.

Methodology: L.S.F., Z.W., D.M., S.V., T.R., B.A.

Investigation: L.S.F., Z.W., D.M., S.V., T.R., B.A.

Visualization: L.S.F., Z.W., D.M., S.V., T.R., B.A.

Funding acquisition: T.P., P.V.

Project administration: L.S.F., T.P.

Supervision: T.P., P.V.

Writing – original draft: L.S.F., Z.W., D.M., T.R., B.A., A.C.

Writing – review & editing: L.S.F., T.R., B.A., A.C., T.P., P.V.

## Data and materials availability

The datasets and scripts used in this manuscript are available at: https://github.com/CUB-IGL/Network-analyses-reveal-global-and-local-glioma-related-decreases-in-ipsilesional-structural-connect

## Supplementary Materials

### Materials and Methods

#### Patient selection

We included n=37 left- and right-handed adult patients in this study (15 females, 22 males, the average age was 48.24, SD = 16.47, age range 20-78). Only patients with an initial diagnosis of unilateral WHO grade II, III & IV gliomas (13 WHO grade II, 10 WHO grade III, 14 WHO grade IV) and without a midline shift in structural images were included (Table 1 & Fig. S1). All tumors were infiltrating M1 and/or showing adjacency to the corticospinal tract (CST), either in the left (n=16) or right (n=21) hemisphere. Patients with recurrent tumors, previous radiochemotherapy or non-glial tumors were not considered.

#### Clinical assessment

We used the National Institutes of Health Stroke Scale (NIHSS) to objectively quantify the impairment caused by the tumor (*16*). The NIHSS includes the areas of level of consciousness, eye movements, integrity of visual fields, facial movements, arm and leg muscle strength, sensation, coordination, language, speech and neglect. Each impairment is scored on an ordinal scale of 0 to 2, 0 to 3, or 0 to 4, with the scores adding up to a total score of 0 to 42 (*16*). Besides NIHSS, we used the British Medical Research Council grade (MRC) to assess motor status, where 0 means no muscle activation and 5 means normal muscle strength (*17*).

#### Navigated TMS

All patients were examined with navigated transcranial magnetic stimulation (nTMS) to perform preoperative functional mapping as described in our previous studies (*18, 19*). nTMS is performed by placing a handheld electromagnetic coil on the subjects’ skull to excite neurons and provoke motor evoked potentials (MEP) which are recorded using a connected electromyography (EMG) unit (*20, 21*). Depending on the pathology’s location the EMG activity of different muscles is measured: commonly utilized muscles are the abductor pollicis brevis, first dorsal interosseus (FDI) and abductor digiti minimi for the upper extremity. When examining the motor function of the lower extremity, commonly used muscles are the tibialis anterior and the abductor hallucis. First, the *hot spot* (*22*) of the FDI muscle was identified by applying TMS in a dense grid and with different coil rotations to achieve the best topographic accuracy (*19*). Then, the resting motor threshold (RMT, in V/m), defined as the lowest stimulation intensity sufficient to induce a MEP in at least 5 out of 10 stimulations (≥ 50 μV), was determined at the top of the cortex for each hemisphere. Peritumoral mapping was then performed for the upper (stimulation intensity: 110% RMT) and lower (median stimulation intensity: 130% RMT) extremities (*23*). Finally, mapping (stimulation intensity: 105% RMT) was performed to specifically outline the primary motor cortex along the precentral gyrus.

#### MRI data acquisition

MRI data were acquired on a Siemens Skyra 3T scanner (Erlangen, Germany) equipped with a 32-channel receiver head coil at Charité University Hospital, Berlin, Department of Neuroradiology. These data consisted of a high-resolution contrast enhanced T1-weighted structural scan (TR/TE/TI 2300/2.32/900 ms, 9° flip angle, 256 × 256 matrix, 1 mm isotropic voxels, 192 slices, acquisition time: 5 min) and a single shell dMRI 2 × 2 × 2 mm^3^ voxels, 128 × 128 matrix, 60 slices, 3 b0 volumes) image data set, acquired at b = 0 and 1000 s/mm^2^ with 5 and 30 volumes respectively, for a total acquisition time of 12 minutes. Additionally, T2-weighted and 3D fluid attenuated inversion recovery (FLAIR) and subtraction sequences were performed.

#### T1-weighted structural MRI preprocessing

All T1-weighted images were registered to the dMRI data sets using the Linear Image Registration Tool (FLIRT) of FMRIB Software Library (FSL, v6.0) (*24*). Before obtaining a whole-brain parcellation scheme, the gliomas were manually segmented using Insight Toolkit (ITK) snap, with additional reference to T2 and FLAIR images (*25*). After tumor segmentation, the Clinical Toolbox for Statistical Parameter Mapping (SPM) was used for enantiomorphic lesion filling (*26, 27*). After that, the structural T1-weighted images were processed using FastSurfer’s (*28*) deep learning-based processing of structural human brain MRI data, replicating FreeSurfer’s anatomical segmentation (*29, 30*). FastSurfer outputs subject-specific anatomical segmentations including surface reconstructions and cortical parcellations, following the Desikan–Killiany–Tourville (DKT) atlas, resulting in 76 gray matter nodes (*31, 32*). All results were visually inspected before subsequent computations. Prior to tractography, the structural T1-weighted images were used to generate a five-tissue-type (5TT) image based on Hybrid Surface and Volume Segmentation (HSVS), by using FastSurfer output and FSL tools (*13, 15, 28, 29, 33*). Following this step, lesion masks of voxels were manually set to the pathological tissue volume fractions in the 5TT images (cf. Fig. S2).

#### Diffusion MRI preprocessing and tractography

The preprocessing of dMRI data included the following and was performed within MRtrix3 (*15*) in order, as described earlier in (*34*): denoising (*35*), removal of Gibbs ringing artefacts (*36*), correction of subject motion (*37*), eddy-currents (*38*) and susceptibility-induced distortions (*39*) in FSL (*33*), and subsequent bias field correction with ANTs N4 (*40*). Each dMRI data set and processing step was visually inspected for outliers and artifacts. We defined a threshold of scans with more than 10% outlier slices due to excessive motion, however, this was not exceeded in any patient. We upsampled the dMRI data to a 1.3 mm isotropic voxel size before computing FODs to increase anatomical contrast and improve downstream tractography results and statistics (*41*). For voxel-wise modelling we used a robust and fully automated and unsupervised method. This method allowed to obtain tissue-specific response functions for white and grey matter and cerebrospinal fluid (CSF) from our data using spherical deconvolution for subsequent use in multi-tissue CSD-based tractography (*42-44*).

#### Streamline tractography

For tractography, two tracking algorithms were used. Probabilistic tractography was performed with the 2nd-order integration over fiber orientation distributions (iFOD2) algorithm and additional usage of the anatomically constrained tractography (ACT) framework using the 5TT image (*12, 13, 15*). Tracking parameters were set to an FOD amplitude cutoff value of 0.06, a streamline minimum length of 5 x voxel size and a maximum streamline length of 250 mm. For each tractogram, we computed 50’000’000 streamlines. Further streamline tractography parameters included backtracking, to allow tracks to be truncated and re-tracked if a poor structural termination was encountered, cropping streamline endpoints as they cross the gray matter-white matter interface and determining seed points dynamically (*45*). Subsequently, the whole-brain tractograms were filtered to 10’000’000 streamlines with spherical-deconvolution informed filtering of tractograms (SIFT) such that the streamline densities matched the FOD lobe integrals (*14, 46*). The lesion masks were used to exclude possible underlying streamlines (cf. Fig. S2). Each tractogram was visually inspected for proper streamlines generation after initial tractogram reconstruction, after SIFT and after exclusion of streamlines underlying the lesion masks.

In addition to probabilistic tractography, we performed deterministic tractography with the SD_STREAM algorithm, which is as well based on FOD input. The ACT framework was employed here as well. 4th-order Runge–Kutta integration was used to eliminate curvature overshoot (*14, 47*). Tracking parameters included a 45-degree angle, a .625 step size, a maximum streamline length of 250 mm and an FOD cutoff value of .06. As above, for each tractogram, we computed 50’000’000 streamlines, which were filtered to 10’000’000 streamlines by SIFT (*14*).

#### Connectome construction

After whole brain tractogram generation and filtering, we obtained connectome matrices by mapping the streamlines based on their assignments to the node-wise endpoints defined in the FastSurfer based Desikan–Killiany–Tourville (DKT) parcellation (*28, 31, 32*). Furthermore, we modified the lookup table (LUT) by adding brainstem and left and right cerebellum labels from Freesurfer’s LUT to FastSurfer’s DKT LUT to obtain nodes (left and right cerebellum and brainstem) in the connection matrix that represent the cerebral base of the primary motor area connectivity. This resulted in a subject-specific weighted, undirected network represented as a symmetric 79 × 79 adjacency matrix. In this matrix, each node is represented by a DKT area and each edge as the node-wise structural connectivity. The metric of connectivity quantified in the connectome matrix is the number of streamlines (*15*). For network analysis, in order to identify structural connectivity changes between the ipsi- and contralesional hemispheres, the whole-brain connectome matrices were split with a MATLAB (R2018b) script into ipsilesional and contralesional matrices representing the two hemispheres separately. This split resulted in a temporary 39 × 39 adjacency matrix and after inclusion of the brainstem in a 40 × 40 adjacency matrix for each hemisphere. Interhemispheric edges were excluded to rule out the confounding effect of the opposing hemisphere in subsequent analyses.

#### Statistical analysis

Statistical analysis was performed by MRtrix3 (*15*) for connectome group-wise statistics at the edge level using non-parametric permutation testing via the threshold-free network-based statistics (TFNBS) algorithm (*48*). The network-based clustering was computed with a default n=5’000 shuffling of data for nonparametric statistical inference. We added the hemispheric tumor position as a covariate to account for hemispheric differences (*49*). TFNBS provides multiple hypothesis testing at the level of interconnected subnetworks and controls family-level errors (FWE) in the performance of analyses associated with a particular effect or contrast of interest (*1, 50*). TFNBS overcomes some of the limitations of the generic procedure (such as the false discovery rate, FDR), which computes statistical tests and the corresponding p-value independently for each link and considers only the strength of that compound. NBS performs a univariate mass testing procedure to identify those connections that exceed a statistical test threshold and belong to a specific connected component. Finally, a corrected p-value is calculated for each component using the null distribution of the maximum size of the connected component, which is derived empirically using a non-parametric permutation method (*1, 50*).

To further characterize the tumor impact on the structural networks, we made use of graph based complex network analyses on the same matrices that were used as input for TFNBS (*2*). We used measures of network efficiency to detect aspects of functional integration and segregation (*2*). We assessed the global efficiency (*51*), a measure of network integration. Global efficiency allows to assess disconnected networks, as paths between disconnected nodes are defined to have infinite length, and correspondingly zero efficiency. Additionally, we measured the local efficiency (*51*), a measure of network segregation. Local efficiency reflects the extent of integration between the immediate neighbors of the given node (*52, 53*). Furthermore, we computed measures of assortativity, degree, centrality, hierarchy, nodal efficiency, and rich club and small world organization to analyze the vulnerability and resilience of the networks and detect possible abnormalities of network connectivity (*2*). These graph theoretical network analyses were performed by GRETNA 2.0.0 (*54*).

To estimate a rank-based measure of association with neuropsychological assessments, we used the FDR-adjusted Spearman rank coefficient within RStudio 1.3.1093 with R version 3.6.3. The plots were generated with the ggplot2 package (*55*).

Furthermore, we performed Pearson correlations to study the relationship between network measures obtained using SD_STREAM- and iFOD2-based tractography. Additional, supplementary statistical plots can be found in Figs. S3-9 and Tables S2-4.

**Figure S1:**
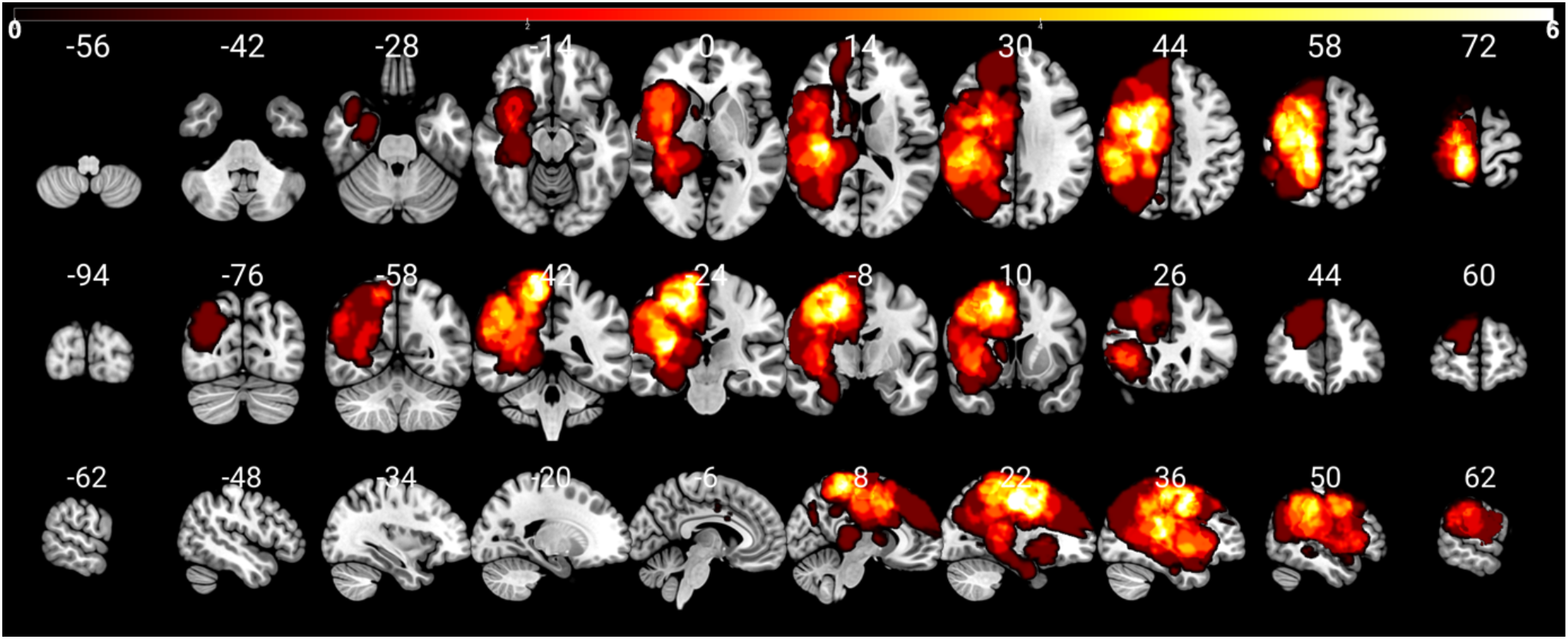
Distribution of the patients’ lesions. The color bar indicates the occurrence of lesions per voxel (white = high quantity, 0-6). To enable a clear comparison of lesion location, lesions of the left hemisphere were mapped to the right hemisphere. The numbers on top of the slices show their position in MNI space.

**Figure S2:**
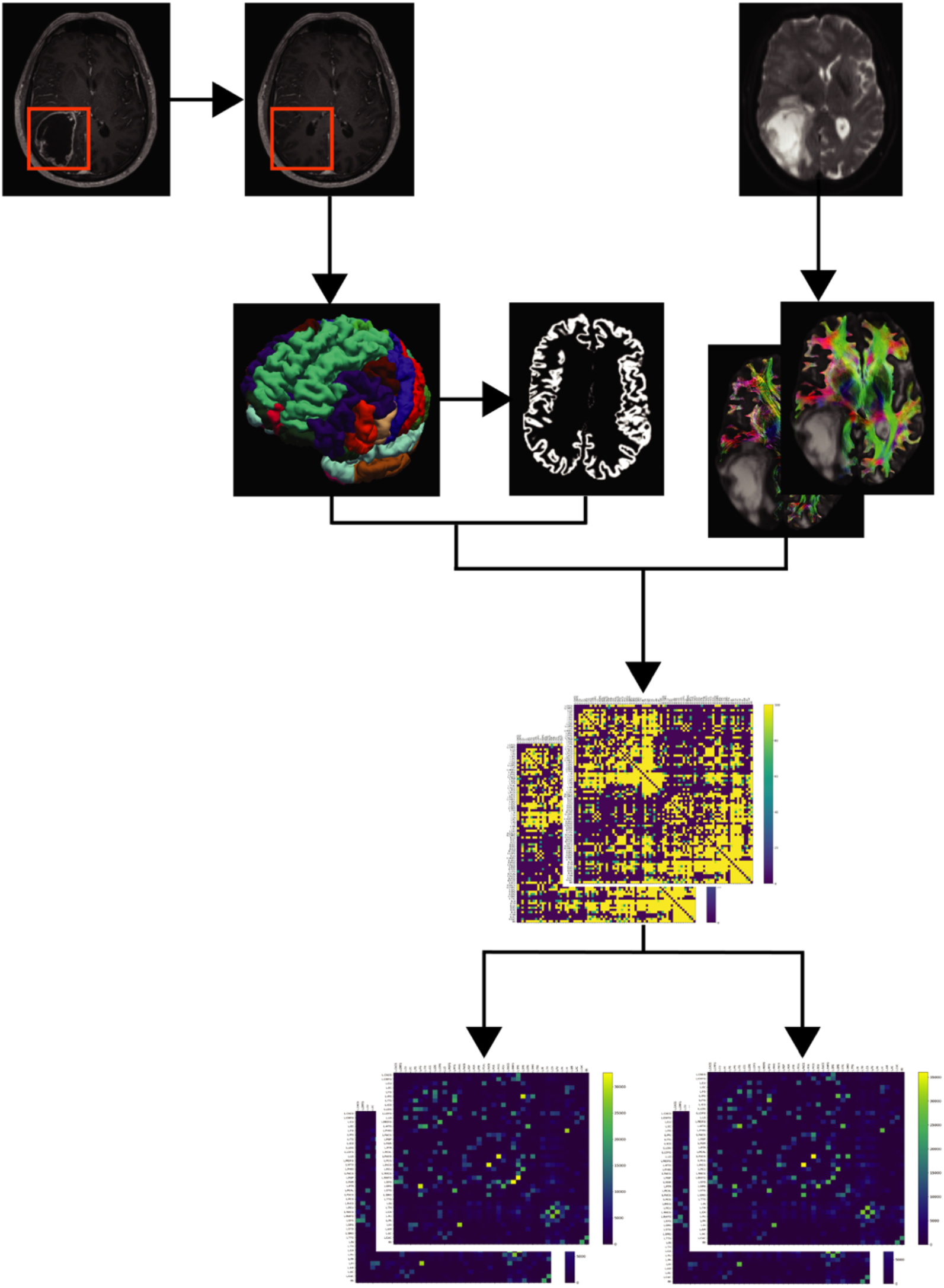
Network reconstruction workflow. Tumors were masked and used for enantiomorphic filling of the T1-weighted structural images (top left), which were then processed by the FastSurfer pipeline to obtain subject-specific parcellations and surface reconstructions (middle left). After whole brain tractography within the ACT framework of 50’000’000 streamlines per subject, tractograms were filtered to 10’000’000 streamlines per subject. Further, symmetric connectome matrices were generated and separated into contra- and ipsilesional matrices (bottom).

**Table S1.**
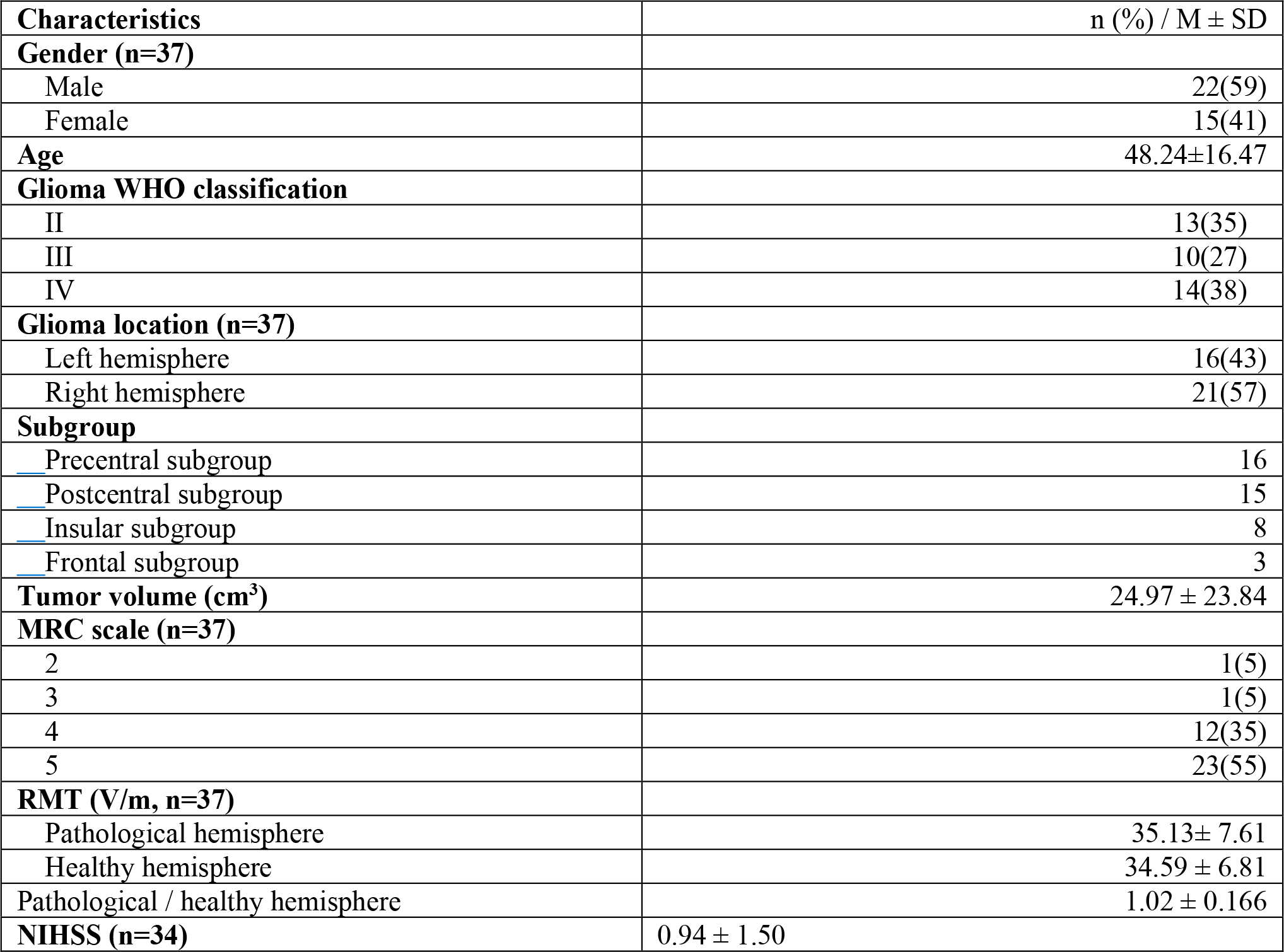
Demographic data and clinical characteristics. MRC scale: Medical Research Council scale, from 0 to 5; RMT: resting motor threshold; NIHSS: National Institutes of Health Stroke Scale.

**Supplementary Fig. S3.**
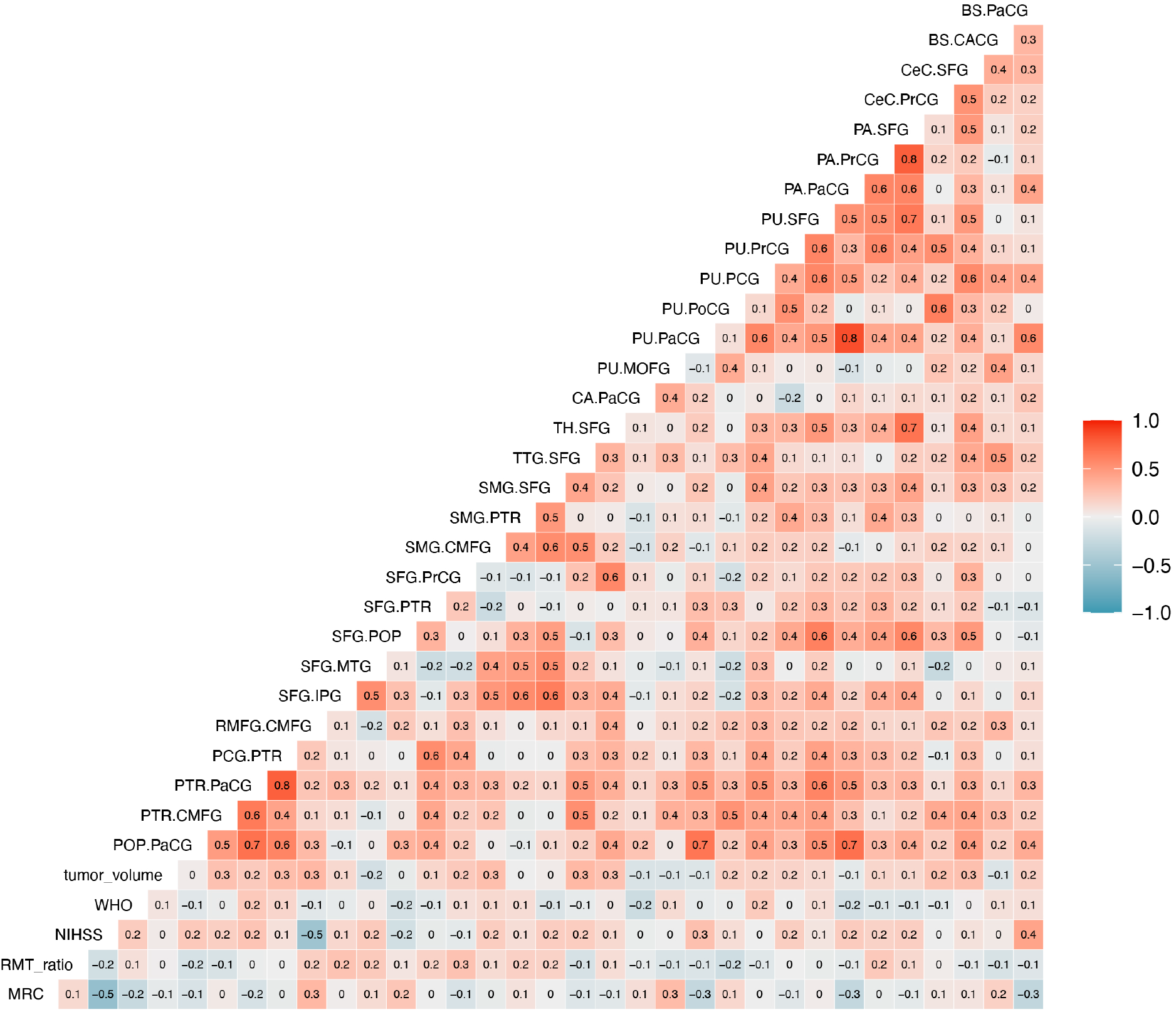
iFOD2 difference of matrices correlogram.

**Supplementary Fig. S4.**
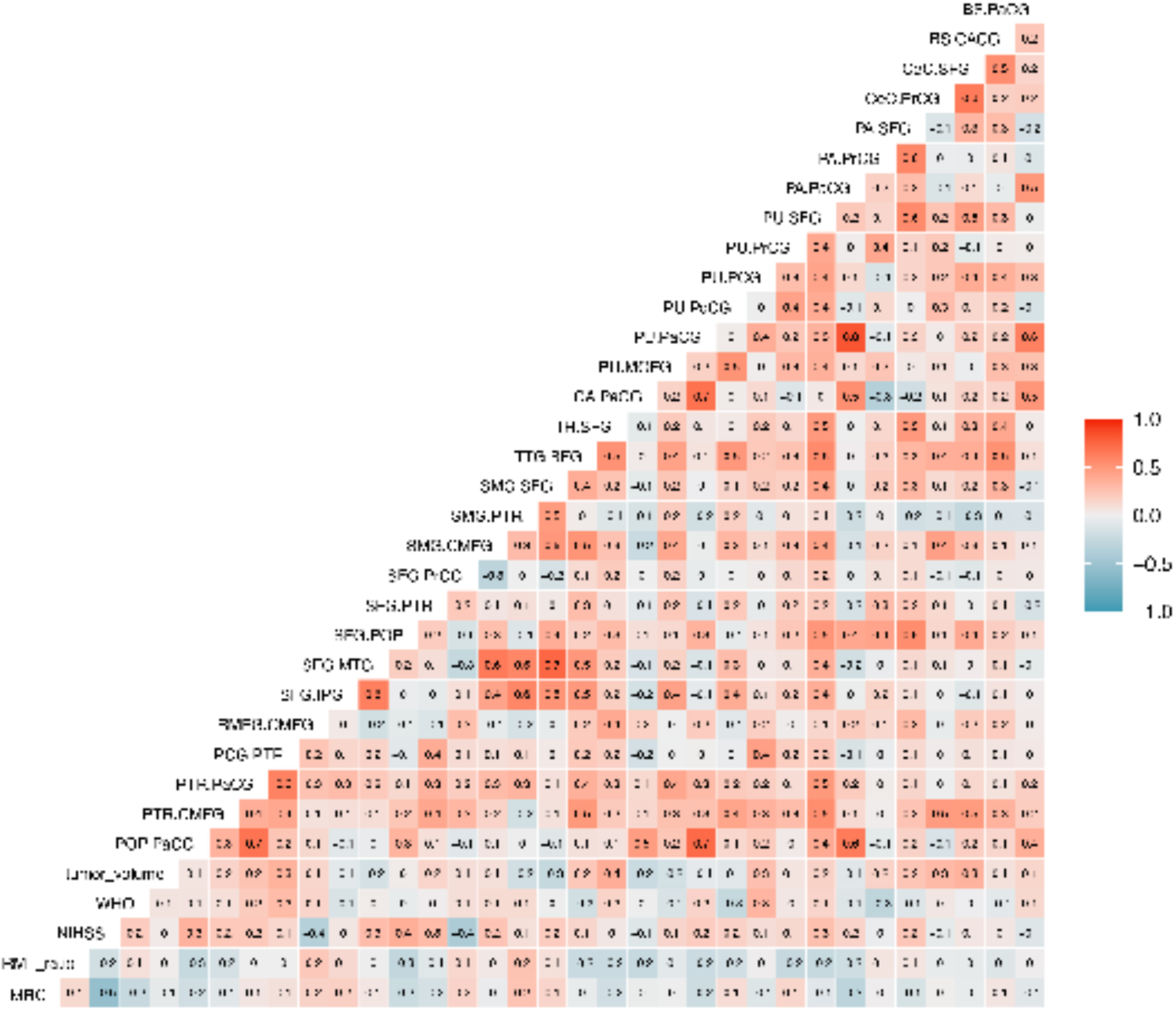
iFOD2 contralesional hemisphere.

**Supplementary Fig. S5.**
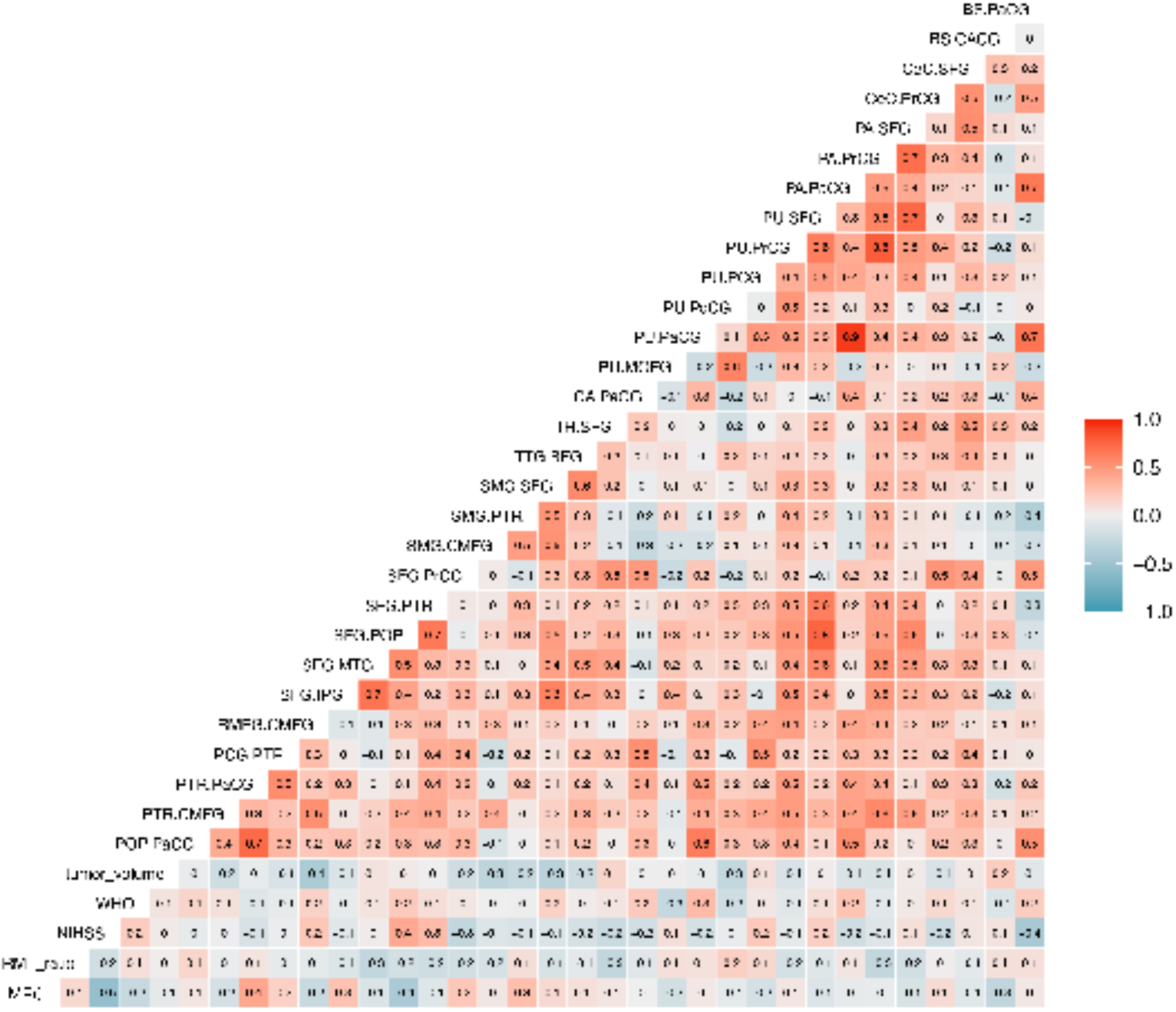
iFOD2 ipsilesional hemisphere.

**Supplementary Fig. S6.**
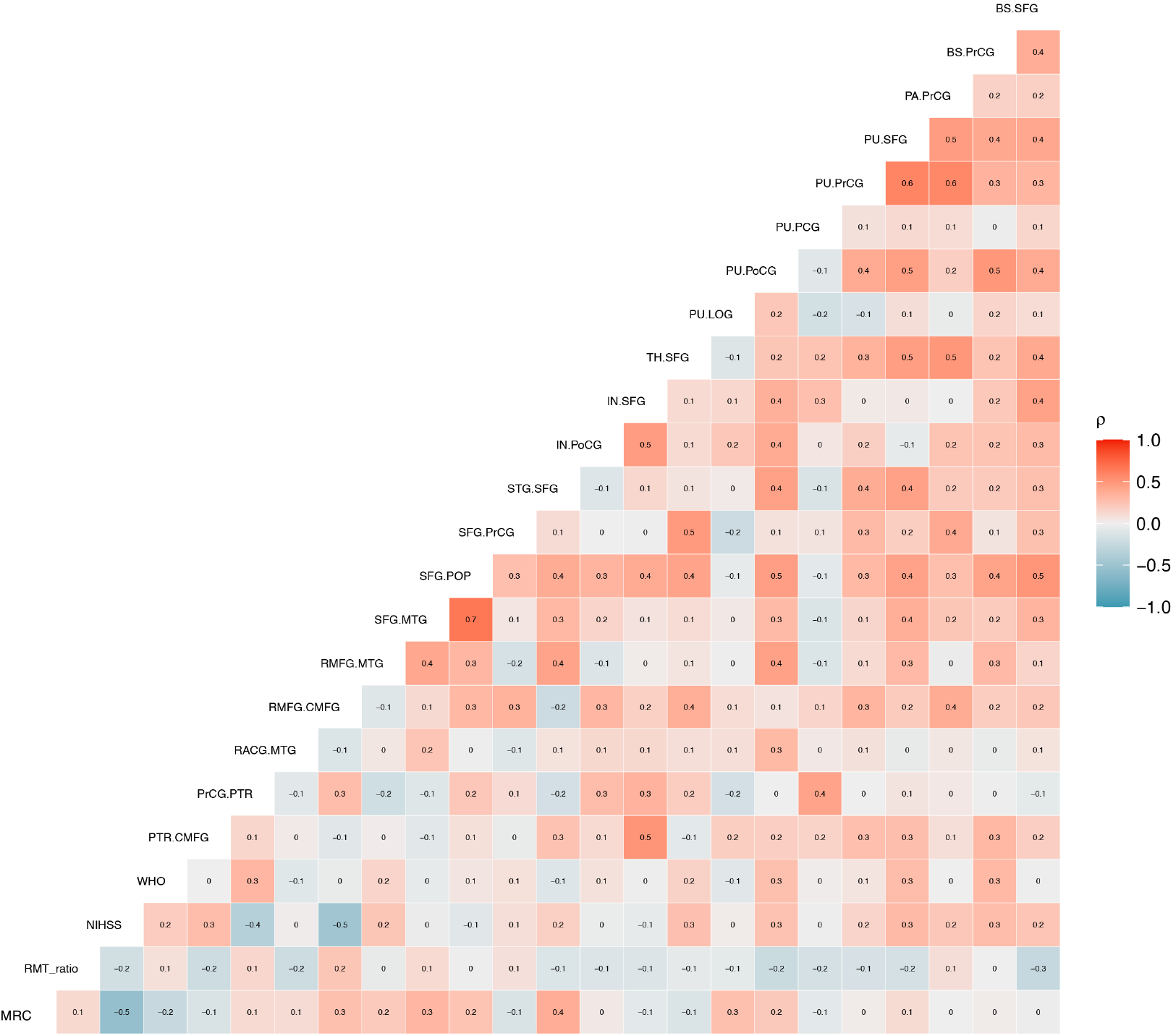
SD_STREAM matrices difference correlogram.

**Supplementary Fig. S7.**
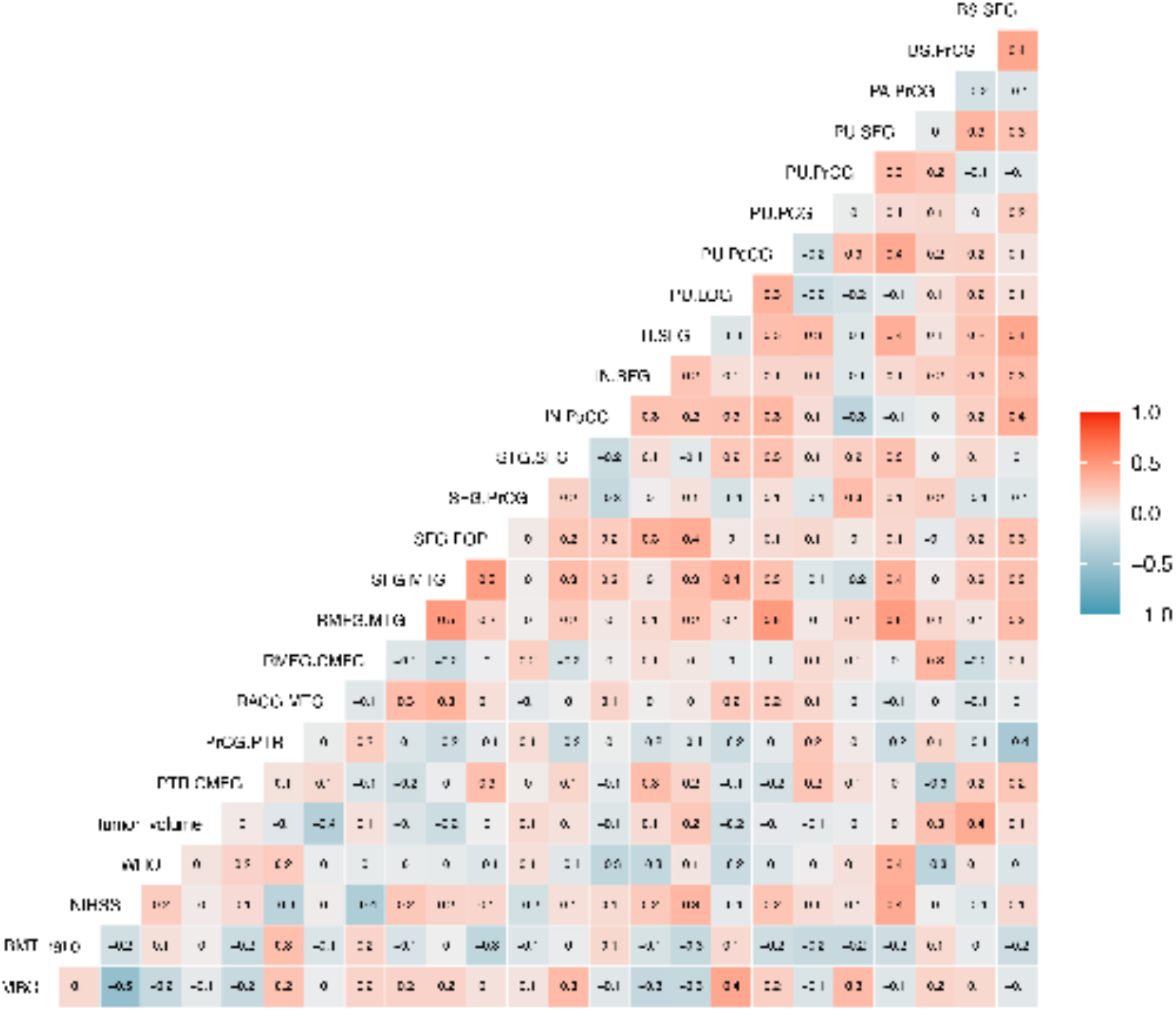
SD_STREAM contralesional hemispheres.

**Supplementary Fig. S8.**
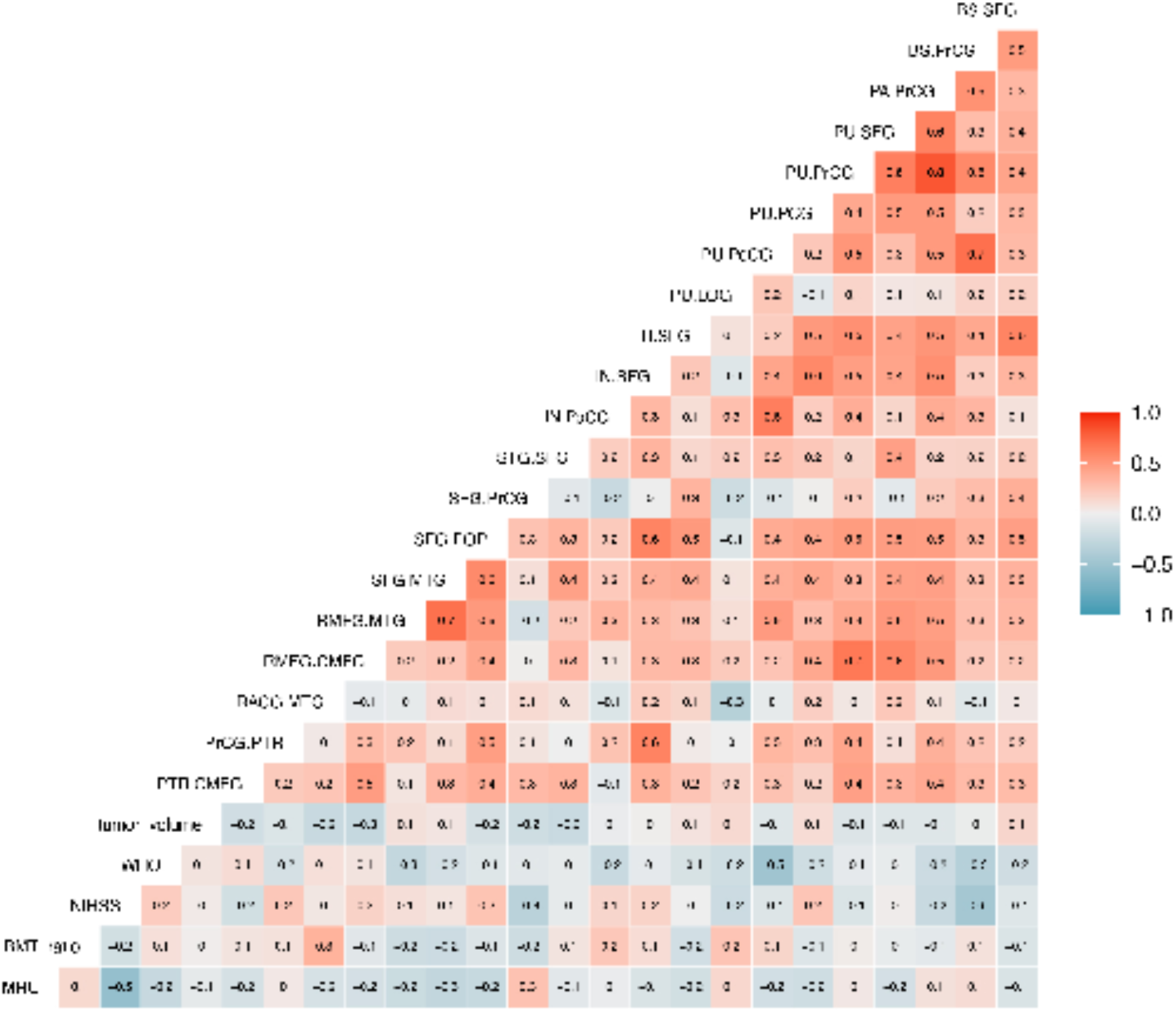
SD_STREAM ipsilesional hemispheres.

**Supplementary Fig. S9.**
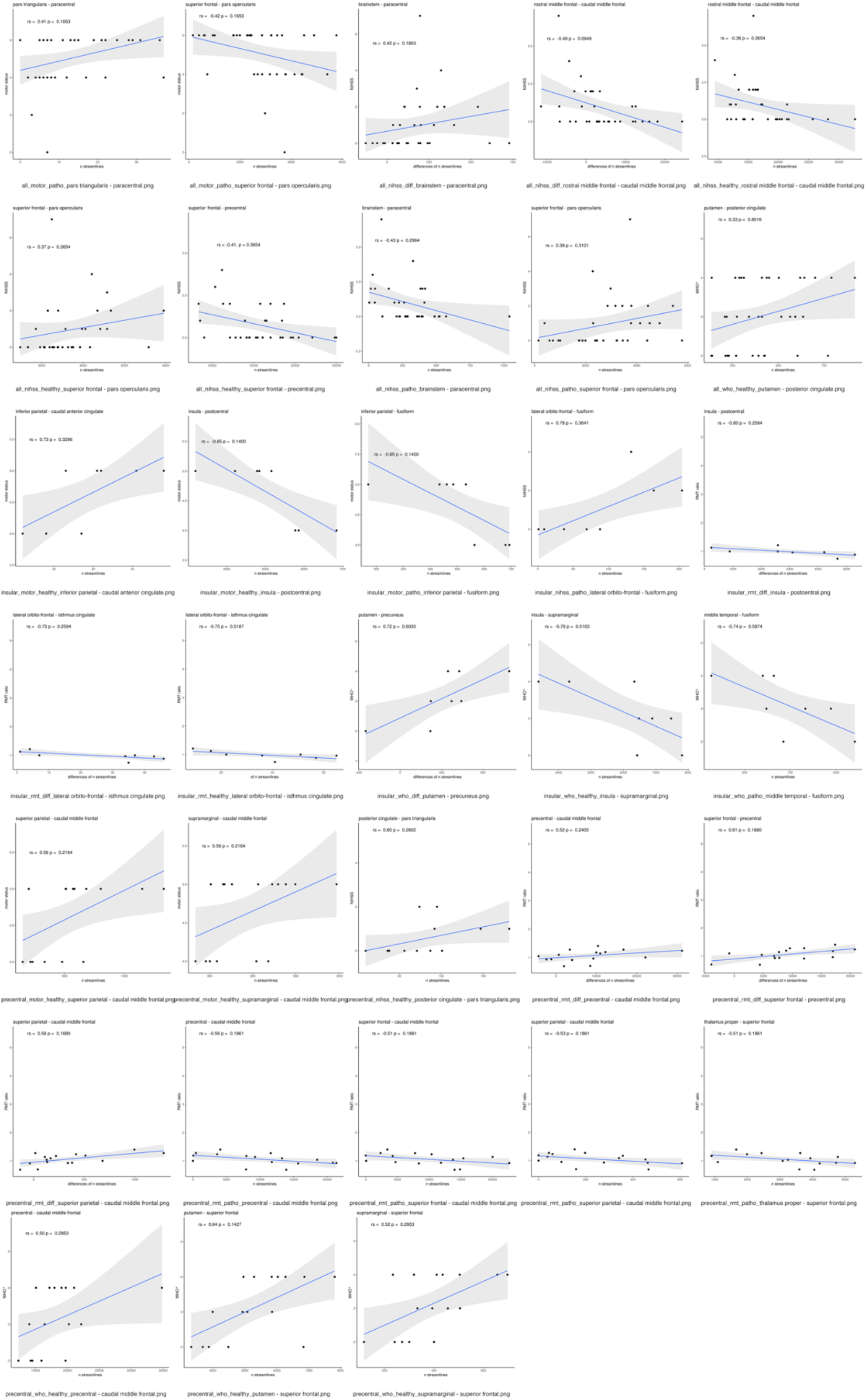
Line plots of FDR-corrected Spearman significant correlations between TFNBS-selected significant edges and NIHSS, RMT ratio, WHO degree and tumor volume.

**Table S2.** iFOD2 significant edges after TFNBS.

| Groups | Edges | FWE adjusted P values |
| --- | --- | --- |
| Entire | POP-PaCG | .0206 |
| Entire | PTR-CMFG | .0006 |
| Entire | PTR-PaCG | .001 |
| Entire | PCG-PTR | .0206 |
| Entire | RMFG-CMFG | .0144 |
| Entire | SFG-IPG | .0144 |
| Entire | SFG-MTG | .0144 |
| Entire | SFG-POP | .0046 |
| Entire | SFG-PTR | .0446 |
| Entire | SFG-PrCG | .0024 |
| Entire | SMG-CMFG | .002 |
| Entire | SMG-PTR | .0206 |
| Entire | SMG-SFG | .001 |
| Entire | TTG-SFG | .0144 |
| Entire | TH-SFG | .0206 |
| Entire | CA-MOFG | .0446 |
| Entire | CA-PaCG | .006 |
| Entire | PU-MOFG | .0206 |
| Entire | PU-PaCG | .006 |
| Entire | PU-PoCG | .0144 |
| Entire | PU-PCG | .0084 |
| Entire | PU-PrCG | .0446 |
| Entire | PU-SFG | .0084 |
| Entire | PA-PaCG | .0144 |
| Entire | PA-PrCG | .0144 |
| Entire | PA-SFG | .0024 |
| Entire | CeC-PrCG | .006 |
| Entire | CeC-SFG | .0144 |
| Entire | BS-CACG | .0446 |
| Entire | BS-PaCG | .0294 |
| Precentral | POP-PaCG | .0062 |
| Precentral | PTR-CMFG | .0226 |
| Precentral | PTR-PaCG | .0008 |
| Precentral | PoCG-CMFG | .0226 |
| Precentral | PCG-POP | .0062 |
| Precentral | PrCG-CMFG | .009 |
| Precentral | RMFG-CMFG | .0182 |
| Precentral | SFG-CMFG | .0056 |
| Precentral | SFG-PrCG | .0026 |
| Precentral | SPG-CMFG | .0226 |
| Precentral | SMG-CMFG | .0126 |
| Precentral | TH-SFG | .0066 |
| Precentral | PU-SFG | .0182 |
| Precentral | CeC-SFG | .0226 |
| Precentral | BS-SFG | .0066 |
| Insular | IPG-FG | .0008 |
| Insular | ITG-FG | .0304 |
| Insular | LOFG-FG | .0118 |
| Insular | LOFG-ICG | .0412 |
| Insular | LOFG-LOG | .0304 |
| Insular | LG-LOFG | .0304 |
| Insular | MTG-FG | .0304 |
| Insular | IN-ICG | .0234 |
| Insular | IN-LOG | .0304 |
| Insular | IN-PoCG | .0192 |
| Insular | IN-SMG | .0038 |
| Insular | PU-PoCG | .0412 |
| Insular | PU-PCU | .0412 |
| Insular | PA-SMG | .0234 |
| Insular | BS-PaCG | .0424 |

**Table S3.** SD_STREAM significant edges after TFNBS.

| Groups | Edges | FWE adjusted P values |
| --- | --- | --- |
| Entire | PTR-CMFG | .0078 |
| Entire | PrCG-PTR | .0126 |
| Entire | RACG-MTG | .0454 |
| Entire | RMFG-CMFG | .0126 |
| Entire | RMFG-MTG | .0064 |
| Entire | SFG-MTG | .0008 |
| Entire | SFG-POP | .0126 |
| Entire | SFG-PrCG | .0006 |
| Entire | STG-SFG | .0302 |
| Entire | IN-PoCG | .003 |
| Entire | IN-SFG | .001 |
| Entire | TH-SFG | .003 |
| Entire | PU-LOG | .0454 |
| Entire | PU-PoCG | .0026 |
| Entire | PU-PrCG | .0126 |
| Entire | PU-SFG | .003 |
| Entire | PA-PrCG | .0008 |
| Entire | BS-PrCG | .0008 |
| Entire | BS-SFG | .001 |
| Precentral | MTG-CMFG | .0022 |
| Precentral | PTR-MTG | .033 |
| Precentral | PoCG-CMFG | .0022 |
| Precentral | PrCG-CMFG | .0092 |
| Precentral | RMFG-CMFG | .0036 |
| Precentral | SFG-CMFG | .001 |
| Precentral | SFG-POP | .0016 |
| Precentral | SFG-PrCG | .0006 |
| Precentral | TH-SFG | .0052 |
| Precentral | CA-PrCG | .0138 |
| Precentral | CA-SFG | .0036 |
| Precentral | PU-SFG | .0022 |
| Precentral | PA-PrCG | .0214 |
| Precentral | BS-SFG | .0022 |
| Insular | IPG-FG | .0164 |
| Insular | LOFG-FG | .0228 |
| Insular | IN-PoCG | .0024 |
| Insular | IN-SMG | .0038 |
| Insular | PU-PoCG | .01 |
| Insular | HI-STG | .027 |

**Table S4.** Overlapping significant edges after TFNBS in SD_STREAM and iFOD2.

| Groups | Edges |
| --- | --- |
| Entire | PTR-CMFG |
| Entire | RMFG-CMFG |
| Entire | SFG-MTG |
| Entire | SFG-POP |
| Entire | SFG-PrCG |
| Entire | TH-SFG |
| Entire | PU-PoCG |
| Entire | PU-PrCG |
| Entire | PU-SFG |
| Entire | PA-PrCG |
| Precentral | PoCG-CMFG |
| Precentral | PrCG-CMFG |
| Precentral | RMFG-CMFG |
| Precentral | SFG-CMFG |
| Precentral | SFG-PrCG |
| Precentral | TH-SFG |
| Precentral | PU-SFG |
| Precentral | BS-SFG |
| Insular | IPG-FG |
| Insular | LOFG-FG |
| Insular | IN-PoCG |
| Insular | IN-SMG |
| Insular | PU-PoCG |

